# Accurate prediction of gain- and loss-of-function variants in GABA_A_ receptors

**DOI:** 10.64898/2025.12.03.25341551

**Authors:** Christian M. Boßelmann, Sebastian Ortiz, Rebekka Staal Dahl, Vivian W. Y. Liao, Serene El-Kamand, Susan X. N. Lin, Anthony Sze Hon Kan, Tobias Brünger, Dennis Lal, Holger Lerche, Jules Kreuer, Nico Pfeifer, Mary Chebib, Nathan L. Absalom, Philip K. Ahring, Rikke S. Møller

## Abstract

GABA_A_ receptors are critical for inhibitory neurotransmission. Variants in genes encoding these receptors are involved in the pathophysiology of both common and rare epilepsy syndromes. Variant effects on channel biophysical function, broadly classified as gain-of-function (GOF) or loss-of-function (LOF), are associated with key clinical characteristics and treatment response. Understanding and predicting variant effects is therefore essential to improve care for individuals with GABA_A_-related disorders. Here, we present GABA_A_ receptor functional variant effect prediction using multi-task phenotypic learning (GENTLY).

We collected clinical data from 505 affected individuals with 272 (likely) pathogenic GABA_A_ receptor variants across *GABRA1, GABRB2, GABRB3*, and *GABRG2*. All variants were evaluated with *in-vitro* electrophysiology using receptor assemblies that reflect heteropentamer composition in heterozygous carriers. Variants were annotated with features based on sequence (e.g. physicochemical properties, conservation), structure (e.g. binding sites, domains), and phenotypes represented by 8185 HPO terms. We trained separate models on all features and without clinical features. Model performance was estimated using ablation, cross-validation, and external validation on a further 197 individuals with 138 (likely) pathogenic GABA_A_ receptors variants.

Our models enable highly accurate prediction of GOF/LOF in GABA_A_ (AU-ROC 0.863-0.946), outperforming state-of-the-art genome-wide predictors (LoGoFunc: AU-ROC 0.495; evo2: AU-ROC 0.559-0.755) and clinical decision-making (decision tree: AU-ROC 0.823). Model scores correlated strongly with GABA sensitivity (*r* = -0.77, *p* < 0.001). Predictions were consistent with expert-based structure-function hypotheses: variants located in transmembrane domains were more likely GOF (*p* < 0.001), and variants in GABA binding sites were more likely LOF (*p* < 0.001). Predictions on variants from population databases behaved as expected: 13,389 population variants were similar to functionally neutral variants, and predictions from (likely) pathogenic ClinVar variants were similar to GOF/LOF variants. Our model may provide additional evidence for 10-29% of 2,295 variants in ClinVar. Lastly, we show that a simple *k*-nearest neighbour algorithm can predict likely clinical characteristics only from variant information (median Lin similarity 0.754 IQR 0.161).

We demonstrate accurate variant effect prediction in GABA_A_ receptors with rigorous validation across the largest dataset of functionally tested variants to date. Our predictions correlate with continuous electrophysiological measurements not directly used during training and conform to known structure-function relationships, supporting their biological plausibility. These predictions may facilitate timely diagnosis, precision treatment, and prognosis of individuals with GABA_A_ receptor related disorders. A web interface, precomputed scores, and ACMG-calibrated score thresholds for all possible variants are openly available.

## Introduction

Variants in genes encoding for GABA type A (GABA_A_) receptor subunits are associated with both common and rare epilepsies, ranging from risk factors for febrile seizures or genetic generalised epilepsy in otherwise healthy individuals to severe developmental and epileptic encephalopathies.^1–6^ GABA_A_ receptors are ligand-gated, primarily postsynaptic chloride channels that mediate inhibitory neurotransmission and thus play a central role in the homeostasis of the excitatory-inhibitory balance.^7,8^ In total, 19 genes encode the GABA_A_ receptor subunits: *GABRA1-6* (α1-6), *GABRB1-3* (β1-3), *GABRG1-3* (γ1-3), *GABRD* (δ), *GABRE* (ɛ), *GABRP* (π), *GABRQ* (θ), and *GABRR1-3* (ρ1-3). Out of these, the following have so far repeatedly been associated with Mendelian disease: *GABRA1, GABRA2, GABRA3, GABRA5, GABRB1, GABRB2, GABRB3, GABRG2, GABRD*.^1,6,7,9–17^

Previously, GABA_A_ receptor variants associated with epilepsy were thought to only lead to loss of receptor function (LOF, e.g. reduced GABA sensitivity and/or decreased GABA-evoked current amplitudes), in turn resulting in impaired inhibitory neurotransmission, neuronal network hyperexcitability, and thus seizures. This assumption was challenged by recent research that demonstrated gain-of-function effects (GOF, primarily observed as increased GABA sensitivity) in variants associated with vigabatrin hypersensitivity and distinct neurodevelopmental phenotypes.^1,12–15,17^ Thus, knowledge of variant functional effect (GOF/LOF) has become critical information for translational care in these often severely affected individuals. In related ion channel and transporter disorders, variant functional effect can facilitate diagnosis, inform genetic counselling, enable candidate precision therapies, and thus improve prognosis.^18–20^

Direct experimental evidence from *in-vitro* electrophysiology remains the gold standard to ascertain variant functional effect. However, this traditional approach represents a potential challenge to translational care as electrophysiology requires considerable time, experience and resources and thus is not available for routine clinical care. With the increasing availability of next-generation sequencing, including population-wide newborn screening programs, an increasing number of individuals are affected by variants of uncertain significance and unknown functional effect.^21–23^ The resulting uncertainty delays a timely diagnosis and adds to the substantial clinical, psychosocial, and emotional burden of the ‘diagnostic odyssey’.^24,25^ This translational gap may be addressed by machine learning algorithms that predict variant effect, which has been successfully demonstrated for voltage-gated ion channels (Na_V_, K_V_, Ca_V_) and NMDA receptors.^26–30^

Three novel insights now uniquely enable the development of such a predictive algorithm specifically for GABA_A_ receptor subunits. First, multi-task learning can make efficient use of gene family information to learn from paralogous variants.^29,31–33^ Second, phenotypic learning integrates harmonised clinical data and leverages known and implicit genotype-phenotype correlations.^12,13,30^ Third, variant distance from pore axis and allosteric pathways separates functional subgroups.^28,32,34^ Here, we integrate these concurrent developments to train our model on the largest clinical and experimental dataset to date. The resulting score, GABA_A_ receptor functional variant effect prediction using multi-task phenotypic learning (GENTLY), outperforms state-of-the-art genome wide predictors and clinical decision-making. We demonstrate biological plausibility by correlating model scores with GABA sensitivity and conforming to expert-based structure-function hypotheses. Lastly, the resulting score has clinical utility by separating case from population variants and allowing for inference of likely clinical characteristics.

## Materials and methods

### Clinical data collection

Individuals carrying (likely) pathogenic variants, classified according to ACMG criteria^35^, in genes encoding GABA_A_ receptor subunits were recruited through an international network of collaborating paediatric and adult neurologists and medical geneticists including the European Reference Network for Rare and Complex Epilepsies (ERN-EpiCARE). Additionally, individuals were recruited via patient advocacy groups (CURE GABA-A). Electronic data capture was standardised with case report forms via REDCap (Research Electronic Data Capture) hosted at the Danish Epilepsy Centre in Dianalund.^36^ Clinical data included age at seizure onset and was further harmonised using the human phenotype ontology (HPO), 2024-12-12 Release.^37^ The study was approved by the local Institutional Review Board (EMN-2024-01998). Data are reported in line with the Strengthening Reporting of Observational Studies in Epidemiology (STROBE) statement.

### Functional experiments

Functional evaluation of variant receptors were performed using a custom made two-electrode voltage clamp apparatus described previously.^1,13,17^ Briefly, concatenated pentameric receptor constructs using human GABA_A_ receptor subunits were used that allowed for the systematic introduction of a point mutated subunit into the first, second, third or fourth position of a γ2-β2-α1-β2-α1 or γ2-β3-α1-β3-α1 construct. Mutant constructs were made and verified by sequencing followed by sub-cloning into the concatenated construct using standard restriction digestion and ligation. Linearised cDNA was generated and cRNA for each concatenated receptor construct was produced using the mMessage mMachine T7 Transcription Kit (Thermo Fisher).

The cRNAs of wildtype and mutant concatenated receptors were injected into oocytes at ∼25 ng cRNA per oocyte. Oocytes were incubated for 2 days at 18°C in modified Barth’s solution (96 mM NaCl, 2 mM KCI, 1 mM MgCl_2_, 1.8 mM CaCl_2_, 5 mM HEPES, 2.5 mM sodium pyruvate, 0.5 mM theophylline, and 100 mg/L gentamicin; pH 7.4). All recordings were performed at room temperature. Oocytes were placed in a recording chamber, and of ND96 solution (96 mM NaCl, 2 mM KCl, 1 mM MgCl_2_, 5 mM HEPES, 1.8 mM CaCl_2_, pH 7.4) was continuously perfused. The pipettes were backfilled with 3 M KCl and had open pipette resistances from 0.4 to 2 MΩ when submerged in OR2 solution. Oocytes were voltage clamped using an Axon GeneClamp 500B amplifier (Molecular Devices) at a holding potential of -60 mV. Amplified currents were low-pass filtered at 20 Hz using a four-pole Bessel filter (Axon GeneClamp 500B), digitized using a Digidata 1322B (Molecular Devices) and sampled at 200 Hz on a personal computer using the pClamp 10.2 suite (Molecular Devices). Episodic traces following triggering events representing responses to individual applications were collected.

On each experimental day, the functional properties of wildtype receptors were assessed along with the mutant receptors to eliminate the impact of inter-day variation and variation between batches of oocytes. To assess maximum current amplitudes, 10 mM GABA was applied. GABA concentration-response relationships were then determined by applications of increasing concentrations of GABA to the oocyte. Final datasets for GABA concentration-response were collected from at least 10 independent experiments performed on at least two different batches of oocytes.

Raw traces were analysed using pClamp 10.2. To determine the EC_50_ values of GABA concentration-response relationships, the Hill equation was fitted to peak GABA-evoked current amplitudes for individual oocytes using GraphPad Prism 10:

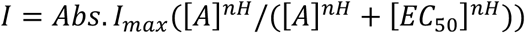

Where Abs. I_max_ is the absolute maximum current, EC_50_ is the concentration that evoke half-maximum response, [A] is the ligand (GABA) concentration and *n*H is the Hill slope. For each individual oocyte, a complete concentration-response curve was recorded as a single determination (n). From the EC_50_value the corresponding log EC_50_ value was calculated. By fitting the Hill equation to all data for each construct, final EC_50_ values were calculated. For each experimental day the mean log EC_50_ for wildtype construct (log EC_50,wt_) was calculated. In addition, the Δ log EC_50_ value for each oocyte containing a mutant construct tested on the same day was calculated using the following equation:

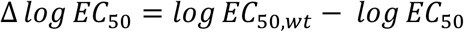

Variants were defined as GOF if the ΔlogEC_50_ was greater than 0.2 and p<0.0001, and LOF where the ΔlogEC_50_ was less than 0.2 and p<0.0001when compared by one-way ANOVA with Dunnett’s post-hoc test.

The normalised maximum GABA-evoked current amplitude (I_max_) was calculated using the peak current evoked by 10 mM GABA at wildtype controls (Abs. I_max,wt_) and mutants (Abs. I_max_) for parallel experiments performed on the same experimental day. To determine the (I_max_) for each individual experiment on a variant following equation was used:

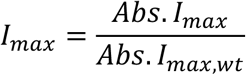

Variants were defined as LOF based on the Imax where the reduction was greater than 50% and p<0.0001when compared with a Mann-Whitney test to wild-type. Thus, each variant had three labels: *(i)* the categorical label of variant effect on channel biophysical function classified as binary gain- or loss-of-function (GOF/LOF); *(ii)* the change in GABA sensitivity, ΔpEC_50_; *(iii)* normalised maximum GABA-evoked current amplitudes.

### Variant annotation

For each gene, variants were described using Matched Annotation from NCBI and EMBL-EBI (MANE) reference transcripts. A list of gene names, transcript IDs, and UniProt accession numbers is provided in Supplementary Table 1. Reference protein sequences were aligned using MUSCLE and used for taxonomy-based multi-task learning as previously described.^38,39^ Protein-level features based on sequence and structure were calculated as previously described, including domain and motif annotation from UniProt under the accession numbers listed above, binding sites, local disorder, conservation, and residue physicochemical properties.^29,30^ Protein-level features were transformed into pairwise similarities with the radial basis function (RBF) kernel 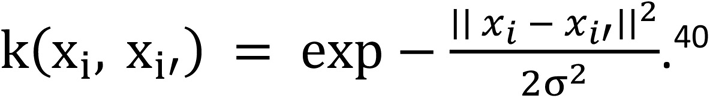^40^

Three-dimensional structural features were calculated using predicted structures from AlphaFold under the UniProt accession numbers listed above.^41^ For each gene, we calculated the Euclidean distance between each pair of residues. Distances were normalised per gene by min-max normalisation. Lastly, distance *d* between each pair of residues *r*_1_ and *r*_2_ was converted to similarity as follows: 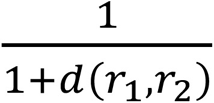 which resulted in a distance-based similarity matrix.

Clinical features were based on HPO terms for each individual and used to calculate pairwise phenotypic similarity as previously described.^30^ If a variant occurred in more than one individual, the union of sets of HPO terms at the variant level was used. The resulting phenotypic similarity matrix was weighed by the absolute difference in age at seizure onset to allow for better representation of age-dependent phenotypes.

### Model development

The model framework used in this study is a multi-task multi-kernel support-vector machine (MTMKL-SVM), which we have previously developed and found to be accurate and robust in the prediction of variants in voltage-gated ion channels.^29,30^ The cost regularization hyperparameter of the support-vector machine was tuned over the range *C* = {10^−6^, 10^−4^, …, 10^6^}. RBF kernel matrices for protein-level sequence and structure features were calculated separately for a range of 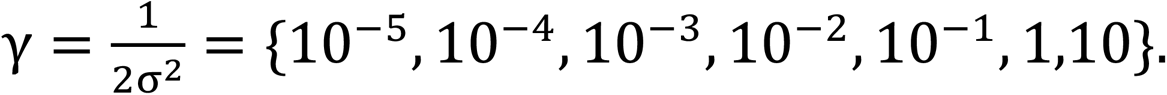 For taxonomy-based multi-task learning, we tested a range of baseline similarities *α* = {1, 2, 3, 5, 10, 100}. For multi-kernel learning, we tested sparse and efficient methods (SimpleMKL, SEMKL)^42–44^ and uniformly weighted kernels.^45^ For calculating phenotypic similarity between sets of HPO terms, we explored different similarity measures: Jaccard, Lin, Resnik, Euclidean, and cosine.^30^ The optimal values for these hyperparameters were estimated using exhaustive grid search (Table 1).

**Table 1.** Model hyperparameters. Abbreviations: MKL – multi-kernel learning; MTL – multi-task learning; SVM – support-vector machine.

| Model | SVM: cost (C) | SVM: gamma ($\gamma$) | MTL: baseline similarity ($\alpha$) | MKL: method | Phenotypic similarity |
| --- | --- | --- | --- | --- | --- |
| Full model | 16 | 0.01 | 100 | Uniform | Lin |
| No clinical features | 4 | 0.01 | 3 | Uniform | – |

Model performance was estimated using five repeats of 5-fold cross-validation. Nested cross-validation was prohibitively time-consuming given the large search space defined above and does not generally result in more precise performance estimates.^46^ Scaling was done on the training splits only to avoid data leakage, and no relevant group imbalance was present. Model predictions obtained from *k*-fold cross-validation (distance from decision boundary) were converted into class (GOF/LOF) probabilities using Platt scaling.^47^ We report the following metrics: *(i)* Accuracy, the number of correct classifications divided by the number of all classifications; *(ii)* Balanced Accuracy, the average recall for each class; *(iii)* Sensitivity (true positive rate, TPR); *(iv)* Specificity (true negative rate, TNR); *(v)* Matthews correlation coefficient (MCC), the correlation coefficient between true and predicted classes; *(vi)* F1 score, the harmonic mean of precision and recall; *(vii)* the area under the receiver operator characteristics (AU-ROC) curve; *(viii)* and the area under the precision-recall curve (AU-PRC).

### Benchmark comparisons

We calculated three state-of-the-art genome-wide functional prediction scores for comparison to our own method. Scores were calculated for all possible non-synonymous single-nucleotide variants across all genes encoding GABA_A_ receptor subunits. First, we implemented LoGoFunc, an ensemble learning method trained on 1492 GOF variants from 344 genes and 13,524 LOF variants from 2030 genes where labels were inferred from literature based on a natural language processing pipeline.^48^ Importantly, the genes in their training dataset included *GABRA1, GABRA3, GABRB2, GABRB3, GABRG2*, and *GABRD*.^48^ This would be expected to positively inflate the performance of LoGoFunc on our benchmark. Next, we implemented the 1 billion and 40 billion parameter models of evo2, a large biological foundation model that can be used to assign delta log–likelihood scores for variants compared to reference sequences (Supplementary Methods).^49^ Score calculation required 4 hours (1.2 kWh) and 67 hours (26.8 kWh) on two NVIDIA H100 Tensor Core GPUs, respectively.

### Population variants

Non-synonymous missense variants in each of the genes encoding GABA_A_ receptor subunits were obtained from two large population databases (gnomAD v4.0 and Regeneron Million Exome Variant Browser; last accessed 12/03/2025) and ClinVar (last accessed 20/11/2025) by querying the gene names listed above while aligning variants to the respective MANE Select reference transcripts.^50–52^

### Phenotype prediction

Relative variant position within the feature space of the model was visualized using UMAP (Uniform Manifold Approximation and Projection) at default parameters, with multivariate normal distributions calculated from the resulting coordinates.^53^ Enrichment of clinical characteristics in functional classes was calculated using two-sided Fisher’s exact test. Likely phenotypes were estimated using a leave-one-out procedure: For each variant, the k-nearest neighbours in the feature space of the model with no clinical features were found. The set of HPO terms of all *k*-nearest neighbours was then compared to the held-out set of HPO terms of the variant of interest using the Lin similarity measure (range 0-1, with higher values denoting higher phenotypic similarity). For comparison, two baseline methods were implemented: First, the set of HPO terms of *k* random variants that were not *k*-nearest neighbours (“Random other”) was compared to the set of HPO terms of the variant of interest. Second, the elements in the set of “Random other” were instead resampled without replacement from all possible terms in the HPO (“Random HPO”). Both baseline estimates were repeated 10-fold.

### Statistical analysis

Continuous values were compared using the Wilcoxon rank-sum test or Student’s t-test, where appropriate based on sample sizes and normality assumptions. Categorical values were compared using Fisher’s exact test. The nominal significance threshold was set at *α* = 0.05. Model decisions were visualized using Cherkassky histograms and calibration plots.^54^ This study was carried out in the R programming language, version 4.4.2, with RStudio, version 2024.9.0.375. Packages used included tidyverse, kernlab, and ontologyX.^55–57^

### Data availability

Model predictions for all possible missense variants are openly available on Zenodo at DOI 10.5281/zenodo.17800749, including annotations of the score ranges and evidence calibration thresholds shown below. An interactive website including our model and cohort summary statistics is openly available (https://nddportals.shinyapps.io/gabaportal/). Raw clinical and experimental data that support the findings of this study are available from the corresponding author upon reasonable request.

## Results

### Accurate and explainable prediction of GABAA receptor subunit variant functional effects

We collected clinical and *in-vitro* electrophysiological data from 505 affected individuals with 272 (likely) pathogenic GABA_A_ receptor variants. 174 variants were unique, and the remaining cases had recurrent variants. Variants were distributed across four genes: *GABRA1* (*n* = 82), *GABRB2* (*n* = 55), *GABRB3* (*n* = 87), and *GABRG2* (*n* = 48). Functional effects were grouped as GOF (*n* = 83), LOF (*n* = 91), neutral effect (*n* = 27), and unknown (untested) effect (*n* = 71). Clinical characteristics were represented with 8185 total and 309 unique HPO terms, with a median of 14 HPO terms per individual (range 5-45; IQR 10-21 terms). Median age at seizure onset was 7 months (range 0-216; IQR 2.5-13 months). Age and number of HPO terms were not correlated (Spearman’s *ρ* = -0.01, *p* > 0.05), suggesting that annotation was not biased towards older individuals.

The full model achieved highly accurate predictions of binary variant effect (GOF/LOF) on internal cross-validation (AU-ROC 0.943 ± 0.035). We conducted ablation studies to gain a better understanding of the model’s behaviour. While removing clinical information deteriorates performance, the overall performance was still competitive (AU-ROC 0.849 ± 0.052). The model without clinical features represents a common use case, e.g. when predicting a variant where clinical information is not yet available or when investigating synthetic variants in scanning mutagenesis. Predictive performance of the full model remained generally robust as other components were removed in turn (Fig. 1A; Table 2; Supplementary Table 2). Interestingly, a model trained exclusively on 3D structural features (pairwise residue distances) was independently predictive (AU-ROC 0.744 ± 0.065) but ablation of the same information from the full model did not deteriorate performance, suggesting that curated structural features (e.g. domains, motifs, ligand binding sites, local disorder) are sufficiently informative.

**Figure 1.**
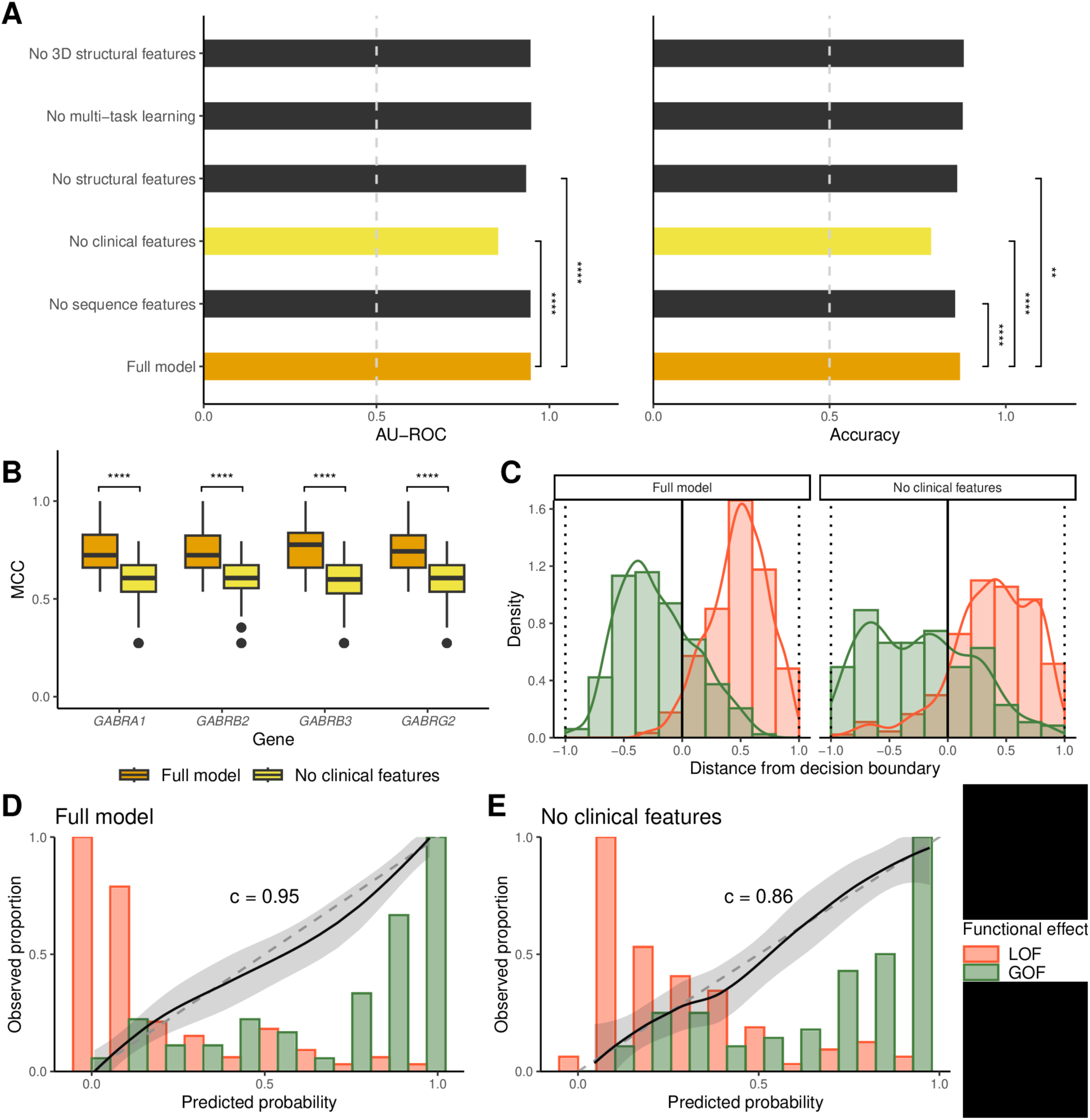
Model performance. **(A)** Model performance estimates (AU-ROC and balanced accuracy) from repeated k-fold cross-validation for the full model and after ablation, where sets of features are removed to investigate feature contribution and model stability. Two-sided Wilcoxon rank-sum test. **(B)** Model performance is stable across tasks (genes) for both the full model and the model with no clinical features. For each task, the full model offers significantly higher performance and consistently achieves a Matthews correlation coefficient (MCC) >0.7, denoting very strong correlation to the experimentally observed effects. Two-sided Wilcoxon rank-sum test. **(C)** Cherkassky histogram of projections for the feature space used by the full model and the model with no clinical features. The bold vertical line at zero distance denotes the SVM hyperplane. **(D-E)** Calibration analysis on cross-validation folds for each model. The dashed diagonal line denotes optimal calibration. Each model was able to discriminate between LOF and GOF variants (c-statistic). Significance: **p < 0.01, \*\*\**p*<0.001, \*\*\*\**p*<0.0001.

**Table 2.**
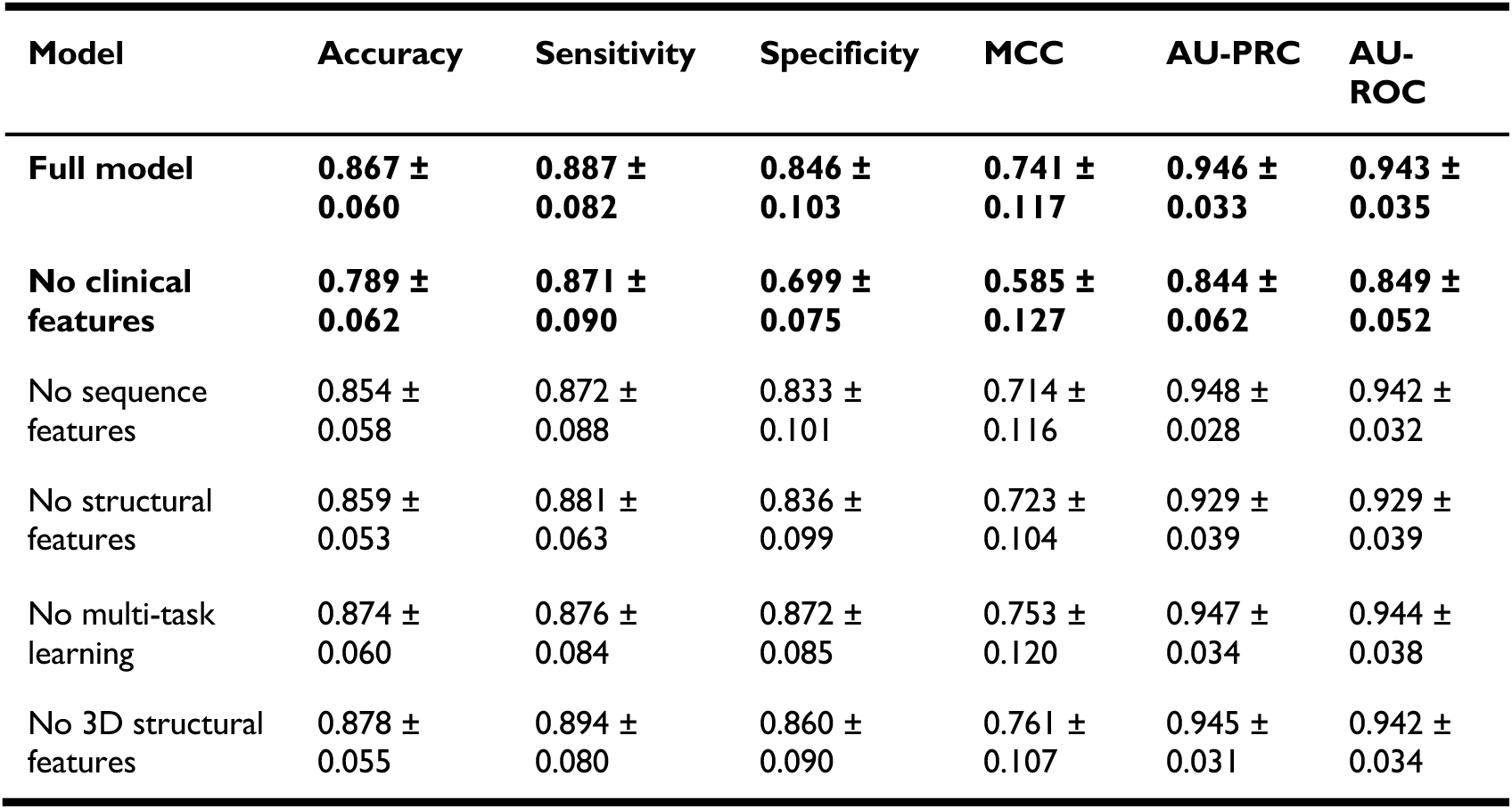
Model performance estimates. Values are shown as mean ± standard deviation. A complete list of performance estimates is provided in Supplementary Table 2. Abbreviations: MCC – Matthews Correlation Coefficient; AU-PRC – area under the precision-recall curve; AU-ROC – area under receiver operating characteristic curve.

| Model | Accuracy | Sensitivity | Specificity | MCC | AU-PRC | AU-ROC |
| --- | --- | --- | --- | --- | --- | --- |
| **Full model** | **$0.867 \pm 0.060$** | **$0.887 \pm 0.082$** | **$0.846 \pm 0.103$** | **$0.741 \pm 0.117$** | **$0.946 \pm 0.033$** | **$0.943 \pm 0.035$** |
| **No clinical features** | **$0.789 \pm 0.062$** | **$0.871 \pm 0.090$** | **$0.699 \pm 0.075$** | **$0.585 \pm 0.127$** | **$0.844 \pm 0.062$** | **$0.849 \pm 0.052$** |
| No sequence features | $0.854 \pm 0.058$ | $0.872 \pm 0.088$ | $0.833 \pm 0.101$ | $0.714 \pm 0.116$ | $0.948 \pm 0.028$ | $0.942 \pm 0.032$ |
| No structural features | $0.859 \pm 0.053$ | $0.881 \pm 0.063$ | $0.836 \pm 0.099$ | $0.723 \pm 0.104$ | $0.929 \pm 0.039$ | $0.929 \pm 0.039$ |
| No multi-task learning | $0.874 \pm 0.060$ | $0.876 \pm 0.084$ | $0.872 \pm 0.085$ | $0.753 \pm 0.120$ | $0.947 \pm 0.034$ | $0.944 \pm 0.038$ |
| No 3D structural features | $0.878 \pm 0.055$ | $0.894 \pm 0.080$ | $0.860 \pm 0.090$ | $0.761 \pm 0.107$ | $0.945 \pm 0.031$ | $0.942 \pm 0.034$ |

Model performance was consistent across each of the four genes (full model: MCC 0.741 ± 0.117, model without clinical features: MCC 0.585 ± 0.127; Fig. 1B). On visual inspection of the histogram of projections (Fig. 1C) and calibration plots (Fig. 1D-E) we found that both models separated well between GOF and LOF variants (c-statistic 0.86-0.95). Lastly, we conducted a sensitivity analysis to test the hypothesis that model performance may be positively inflated by paralogous variants which strongly inform pathogenicity, phenotype and function and are independently predictive.^27,32,33^ We repeated the internal cross-validation, but removed each variant from the test fold located at the same position on the channel family multiple-sequence alignment as any variant in the training fold. This did not significantly impact the performance estimates for our full model (without paralogous variants: AU-ROC 0.939 ± 0.091, two-sided Wilcoxon rank-sum test: *p* = 0.746) and the model without clinical features (without paralogous variants: AU-ROC 0.815 ± 0.054, two-sided Wilcoxon rank-sum test: *p* = 0.421). Thus, our sensitivity analysis demonstrated that our model was not simply memorizing position-specific effects.

Next, we compared model performance to three state-of-the-art genome-wide models that predict variant functional effect: LoGoFunc, evo2-1B, and evo2-40B.^48,58^ For internal validation, we compared prediction scores from *k*-fold cross-validation across the 174 unique variants in our training dataset (Supplementary Table 3). For evo2, the intended use case is zero-shot variant effect prediction. To enable a fairer comparison, we also used evo2 scores to train a supervised model on the same cross-validation folds (Supplementary Methods). This approach to evo2 resulted in moderately improved performance (AU-ROC: 0.756 ± 0.191, accuracy 0.667 ± 0.170; Supplementary Fig. 1) which was still inferior to that of our model (AU-ROC 0.943 ± 0.035, accuracy 0.867 ± 0.060).

**Figure 2.**
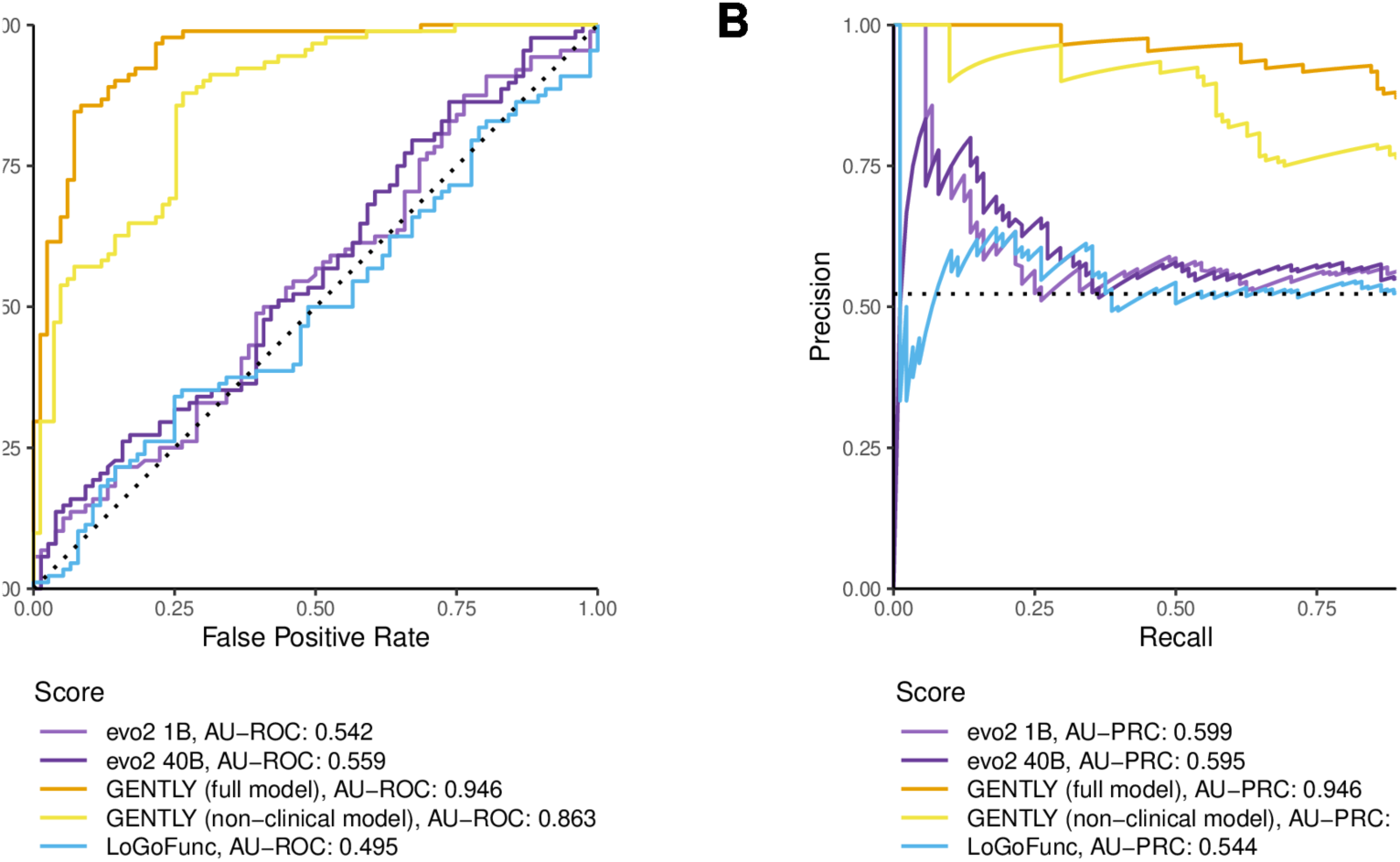
GENTLY outperforms state-of-the-art genome-wide models. **(A-B)** ROC and precision-recall curves for the full and non-clinical models compared to other functional effect prediction models (LoGoFunc, evo2-1B, evo2-40B).

As an additional baseline, we compared our model to clinical decision-making. We fit a decision tree model trained on age at onset and clinical characteristics, mirroring the approach of a clinician experienced in GABA_A_-related disorders. This resulted in a simple six-step model that was surprisingly accurate (sensitivity 0.711 ± 0.112, specificity 0.857 ± 0.084, accuracy 0.787 ± 0.052, AU-ROC 0.823 ± 0.072, AU-PRC 0.848 ± 0.066). The model used a cut-off for age at seizure onset (6 months) and the presence or absence of known genotype-phenotype correlations such as microcephaly, febrile seizures, abnormalities of movement (incl. myoclonus and hyperkinetic movement disorders), focal or generalised seizures (Supplementary Fig. 2).^12–14^ Both of our models still perform better, but this demonstrates that clinical judgment remains highly informative. The primary advantage of our full model including clinical characteristics is ease of use for clinicians who may not be experts in GABA_A_-related disorders.

For external validation, we collected data from 197 individuals with 138 GABA_A_ receptors variants, of which 27 unique variants resulted in GOF/LOF. This small external validation set is not as statistically robust as the internal cross-validation procedure shown above but may serve as an example. Overall, 25 variants were GOF and two variants were LOF. Perhaps most interestingly, this external validation dataset included 23/27 (85%) variants in genes that were not seen by the model during training, so called zero-shot learning (*GABRA3, GABRA5, GABRB1, GABRD*) – a highly challenging prediction problem for the model. Both of our models correctly ranked all four variants in the training distribution and still performed well even on variants in genes not seen during training (full model: AU-ROC 0.818, balanced accuracy 0.841; model with no clinical features: AU-ROC 0.727, balanced accuracy 0.864). Errors were primarily made on variants within two genes in the zero-shot learning group (*GABRA3, GABRD*; Supplementary Table 4). We note that 24/27 (89%) of variants in the external validation dataset were paralogous to variants in our training data. However, the sensitivity analysis we conducted above did not suggest that this would bias the performance estimate. Thus, we demonstrated proof-of-principle that information is transferable across the larger heterogeneous gene family, enabling predictions for new and emerging GABA_A_-related disorders.

Some clinical characteristics in GABA_A_-related disorders may not be seen until a certain age. For example, febrile seizures would be seen around six months, and autism screening is usually conducted at 18-24 months of age. Therefore, not all clinical information may be available when the model is used, and clinicians may wonder how this impacts expected performance. We simulated incomplete observations by removing each HPO term in turn from variants in the training dataset and noting how the prediction confidence changed. For example, not observing microcephaly biased observations towards LOF (mean prediction -0.096) while not observing fever-related HPO terms biased observations towards GOF (mean prediction +0.052). Full results are shown in Supplementary Fig. 3. Overall, these biases were minor given the full score range (-2.32 to 1.74). Indeed, we have previously demonstrated that this model framework is robust to phenotypic noise or sparsity.^30^ Thus, our model may be used for variant interpretation in early life.

### Model predictions are biologically plausible

For further validation, we compared our model predictions to continuous experimental labels: the change in GABA sensitivity compared to wild-type receptors (ΔpEC50) and the normalised maximum current. Information from these measurements were used to assign the label (GOF/LOF) during experimental characterization (Methods). However, we hypothesised that prediction confidence would further correlate with the severity of change in sensitivity to GABA or maximum current beyond simple cut-offs. This would also allow for semi-quantitative assessment of variant functional effect, beyond the binary classification provided by previous models.^26,29,30^ Indeed, we found that scores correlated strongly with sensitivity to GABA (Fig. 3A; Pearson’s correlation coefficient *r* = 0.66-0.77; *p* < 0.001) and moderately with maximum current (Fig. 3B; *r* = 0.38-0.41; *p* < 0.05) in the test dataset from cross-validation. Thus, variants that are more confidently predicted as GOF/LOF cause more severe electrophysiological changes.

**Figure 3.**
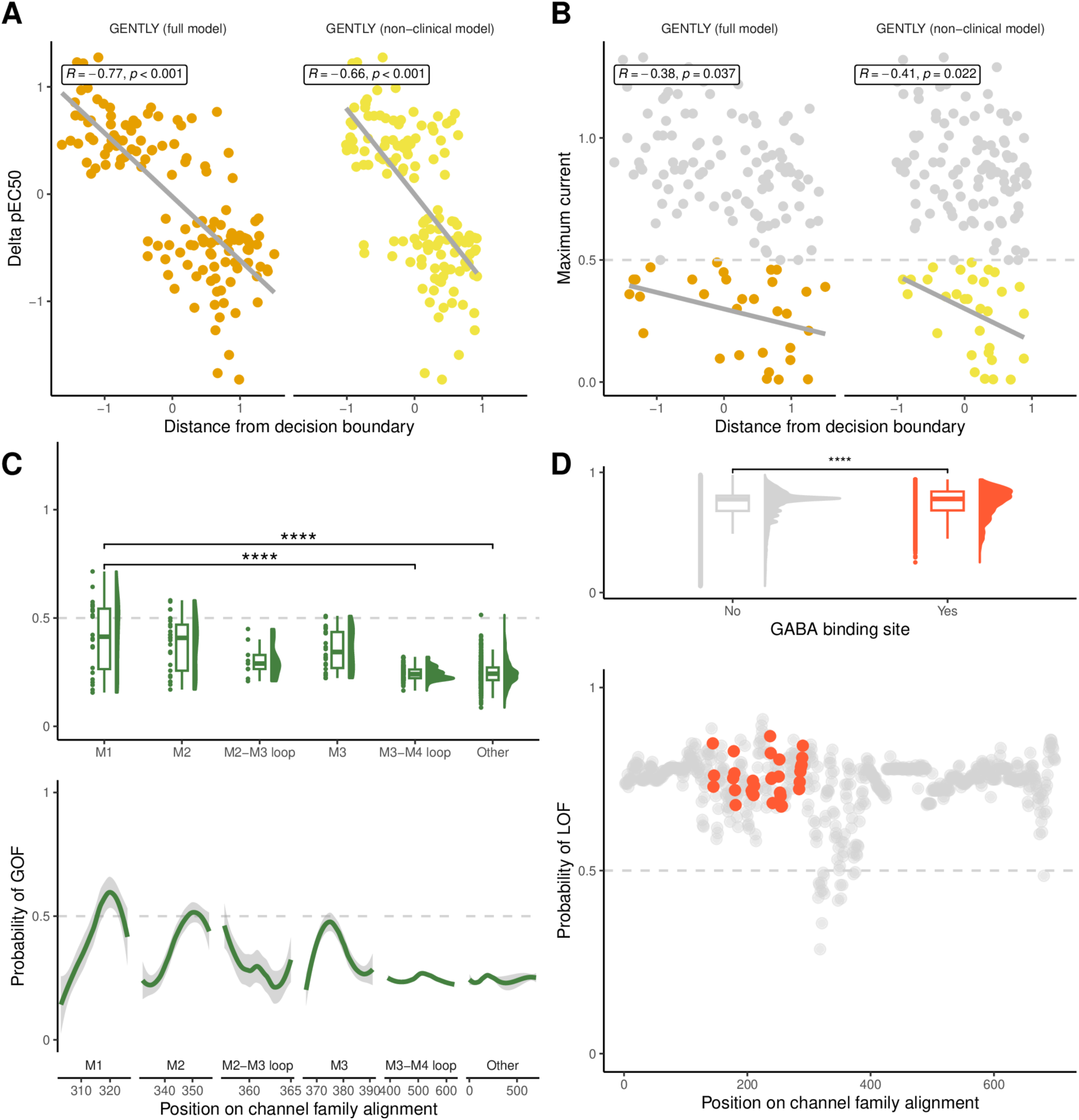
Model predictions are biologically plausible. **(A-B)** Predicted scores from the cross-validation procedure, shown as distance from the decision boundary (i.e. the raw classification used by the SVM model), correlate with continuous electrophysiological measurements (normalised sensitivity to GABA, EC50; normalised maximum current) that were not directly used during model training. Pearson correlation coefficients for both the full model and the model with no clinical features are shown separately. Normalised maximum current values above >0.5 (dashed line) are not meaningfully different from wild-type. **(C)** Model predictions correspond to expert-based hypotheses on structure-function relationships. Prediction scores for all possible GABA_A_ missense variants, averaged for each residue position, were compared across functionally relevant domains. Two-sided t-test. **(D)** Variants at GABA binding sites are significantly more likely to be predicted as LOF by the model. Two-sided t-test. Significance: \*\*\*\**p*<0.0001.

Next, we investigated whether our model conforms to expected structure-function relationships. This would offer additional evidence that our model has learnt biologically plausible information. For this, expert biophysicists (PKA, NLA) prespecified two hypotheses based on literature.^14^ The first hypothesis was that variants at GABA binding sites are more likely to be LOF. Second, variants within the transmembrane segments M1 and M2 are more likely to be GOF than those in extracellular coupling loops, ∼25% of which would be expected to be GOF. For this analysis, we calculated predictions for all possible missense variants in GABA_A_ receptor subunits averaged for each position on the channel family alignment, to allow for comprehensive assessment beyond the positions covered by the training data. Variants within the training data and all paralogous variants were removed for this analysis. We found that variants at GABA binding sites were more likely to have a higher LOF score in our model (two-tailed t-test; mean 0.740 vs. 0.753, *t*(10715): -16.20, *p* < 0.001; Fig. 3C), and accordingly were enriched for variants predicted as LOF (Fisher’s exact test; OR 1.50, 95% CI: 1.36-1.66, *p* < 0.001). Similarly, variants within M1 and M2 were enriched for variants predicted as GOF compared to coupling loops (Fisher’s exact test; OR 6.73, 95% CI: 6.09-7.45, *p* < 0.001; Fig. 3D). Thus, our model successfully learnt expert-level structure-function knowledge.

### Model predictions are clinically meaningful

Future clinical implementation of functional prediction algorithms requires better understanding how these models perform on variants outside of the training distribution. Previous models work under the assumption that the variant of interest has already been classified as (likely) pathogenic according to ACMG criteria, but variant interpretation in rare genetic epilepsies is challenging and prone to misclassification errors.^59–61^ Thus, users may try to predict functional effects of variants that are not disease-related in the individual. Therefore, we investigated how our model behaves when provided with presumably neutral variants from large population databases (gnomAD v4.0, RGC Million Exome Variant Browser). We found that mean scores were different for population variants compared to (likely) pathogenic variants from our training dataset (Fig. 4A) but were similar when compared to 27 variants with neutral functional effect (Fig. 4B). This finding was independently validated using ClinVar, where we found that the score distribution of (likely) pathogenic variant was similar to that of variants from our training data, while the score distribution for variants of uncertain significance and (likely) benign variants was more similar to that of population variants (Fig. 4C). Most population variants fall within the 4th to 7th scores deciles (probability of GOF: 0.208<x<0.283; probability of LOF: 0.717<x<0.792) which provides narrow bounds within which predicted variants should be flagged as potential population or neutral variants, prompting variant reassessment (Fig. 4D).

**Figure 4.**
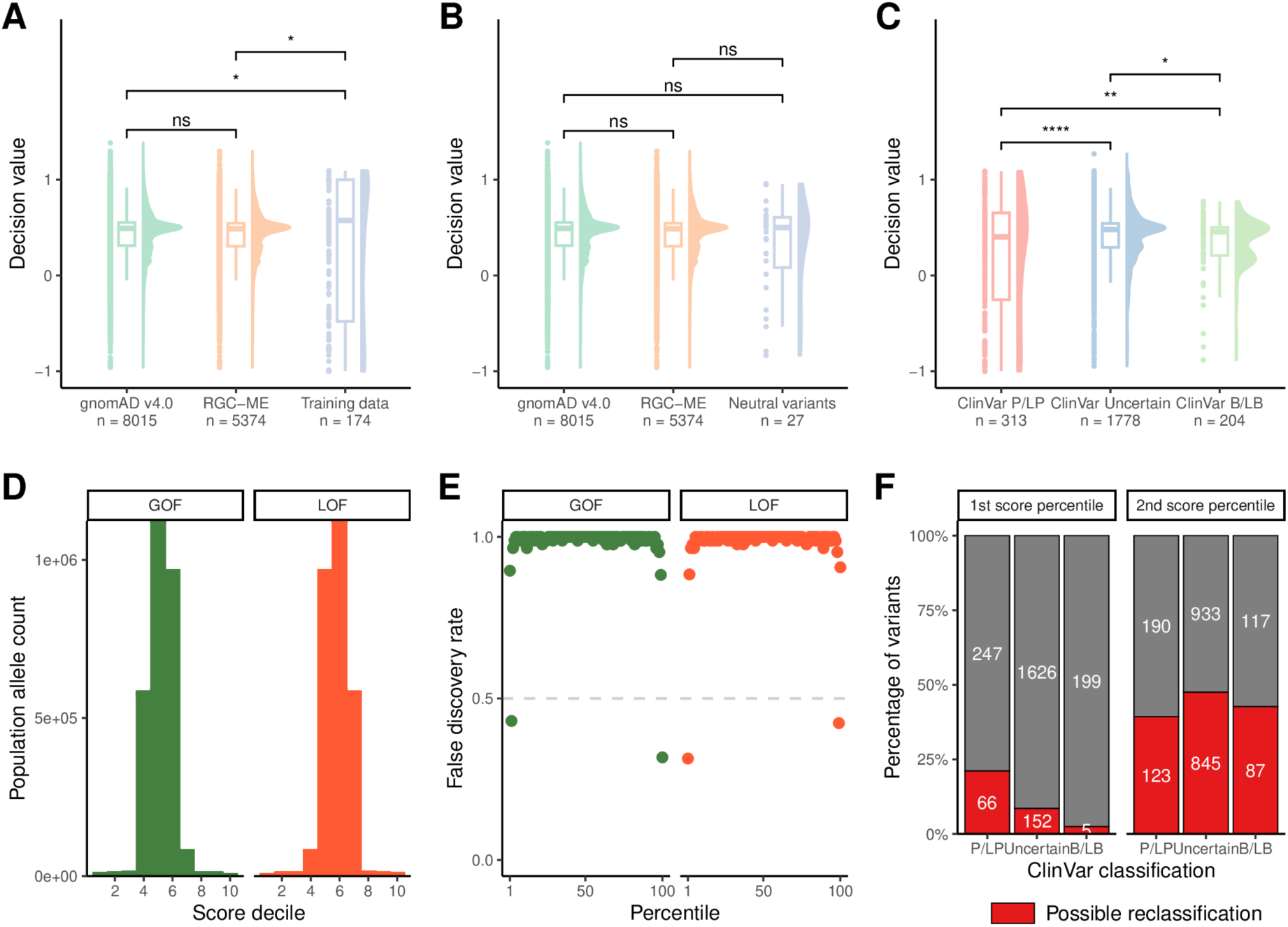
Model predictions are clinically meaningful. **(A)** Model predictions, shown as distance from the decision boundary, are similar across two sets of population variants (gnomAD V4.1.0, RGC Million Exome Variant Browser) but distinct from training variants with functional effects (GOF/LOF). Two-sided t-test. **(B)** Model predictions are not significantly different for population variants and variants with no functional effect (neutral). Two-sided t-test. **(C)** Pathogenic or likely pathogenic (P/LP) variants from ClinVar have scores similar to functional variants, while benign or likely benign (B/LB) variants fall within the score interval of population variants. Two-sided t-test. **(D)** The majority of population variants fall within clearly defined score deciles. **(E)** Variants within the top score percentile can be separated from population variants. **(F)** Variants within the top score percentile may provide additional reclassification evidence for ClinVar variants. Variants observed in the training data were removed for this analysis. Significance: ns – not significant, *p < 0.05, **p < 0.01.

This naturally raises the question whether predictions from our model can be used to distinguish population from pathogenic variants, a task broadly similar to that of pathogenicity prediction algorithms. Variants within the top score percentile (probability of LOF: >0.928; probability of GOF >0.850) were much more likely to be pathogenic variants (Fisher’s exact test; OR 153, 95% CI: 92-260, *p* < 0.001) while variants within the top two score percentiles (probability of LOF: >0.920; probability of GOF >0.729) were still strongly enriched for pathogenic variants (Fisher’s exact test; OR 6.74, 95% CI: 3.05-13.41, *p* < 0.001). Due to the high number of population variants (*n* = 13,389), the false-discovery rate (FDR) for the top score percentile was still 31.4% (Fig. 4E). However, these calculations were done under strict assumptions where any variant irrespective of population allele count (AC) was counted. When this assumption was relaxed (e.g. AC>5), given the role of GABA_A_ receptor variants in common epilepsies and mild phenotypes^62^, the expected performance improved (FDR 4.8%; Supplementary Table 5). Thus, our model can provide complementary evidence when assessing variant pathogenicity but should not be the only source of evidence.

Given the high accuracy of our functional predictions and separability from population variants, our model may be able to provide additional information for variant assessment. We obtained predictions for 2,380 variants from ClinVar, of which 2,295 variants were not observed in our training data. Again, we chose strict assumptions and restricted our analysis to the top two score percentiles defined above. Under conservative estimates, our model can provide additional *in silico* evidence ranging from 152/1778 (9%) to 15/51 (48%) of variants of uncertain significance depending on choice of score threshold. This additional evidence may prompt variant reassessment and reclassification, potentially resulting in more timely and accurate genetic diagnoses. To facilitate use in weighted evidence frameworks, we also calculated score thresholds calibrated to ACMG evidence strengths using a likelihood ratio-based approach (Supplementary Methods, Supplementary Fig. 4, Supplementary Table 6).^63^

We have previously raised the issue that variants in rare genetic epilepsies are often misclassified and highlighted how our model output can be interpreted using careful thresholds to reduce the risk of misclassification and thus the consequences of model misuse. This naturally raises the question of why we chose to directly design a functional prediction model, without gene-specific pathogenicity prediction. Pathogenicity classification goes far beyond the direct use of *in silico* scores, which play only a limited role in the ACMG classification framework.^35^ Further, there already are highly accurate proteome-wide pathogenicity predictors which we have recently benchmarked in ion channel and transporter disorders.^59^ We investigated their accuracy in our dataset, comparing 174 (likely) pathogenic variants from our training data to 2,591 variants from population databases (gnomAD v.4.10, RGC-ME; AC>3). We found that AlphaMissense (AU-ROC 0.905) and REVEL (AU-ROC 0.864) performed well, and REVEL correctly classified variants in our training data according to calibrated ACMG evidence strengths.^63,64^ Variants from ClinVar were not used to avoid circular reasoning from training data of each pathogenicity prediction algorithm.^59^ Thus, currently available pathogenicity prediction models are sufficiently useful for variants in genes encoding GABA_A_ receptor subunits (Supplementary Fig. 5), while our model allows for LOF/GOF prediction.

### Predicting clinical characteristics

It would be clinically useful to directly predict clinical characteristics from variant information, similar to the *SCN1A*-Epilepsy Prediction Model which accurately distinguishes between mild and severe *SCN1A*-associated phenotypes and thus informs prognostic counselling and early-life decisions.^65^ We noted that our model representation already included the same information used for this clinical prediction approach: Clinical characteristics were weighed by age at seizure onset and integrated with variant-level structural information to learn a shared representation which separates between functional subgroups (Fig. 5A-B). These functional subgroups were enriched for distinct clinical characteristics including febrile seizures (*GABRB3* LOF; Fisher’s exact test; OR 18.37, 95% CI: 2.39-838, *p* < 0.001), microcephaly (*GABRB3* GOF; Fisher’s exact test; OR 0.06, 95% CI: 0.01-0.51, p < 0.001), and dystonia (*GABRB3* GOF; Fisher’s exact test; OR 0.09, 95% CI: 0.01-0.73, p < 0.05), as well as other higher-order HPO terms (Fig. 5C). These findings both recapitulate and extend the phenotypic spectrum of associated neurodevelopmental disorders.^14^ An overview across 1,072 HPO terms, of which 108 terms had nominally significant gene-function-phenotype associations, is provided in Supplementary Table 7.

**Figure 5.**
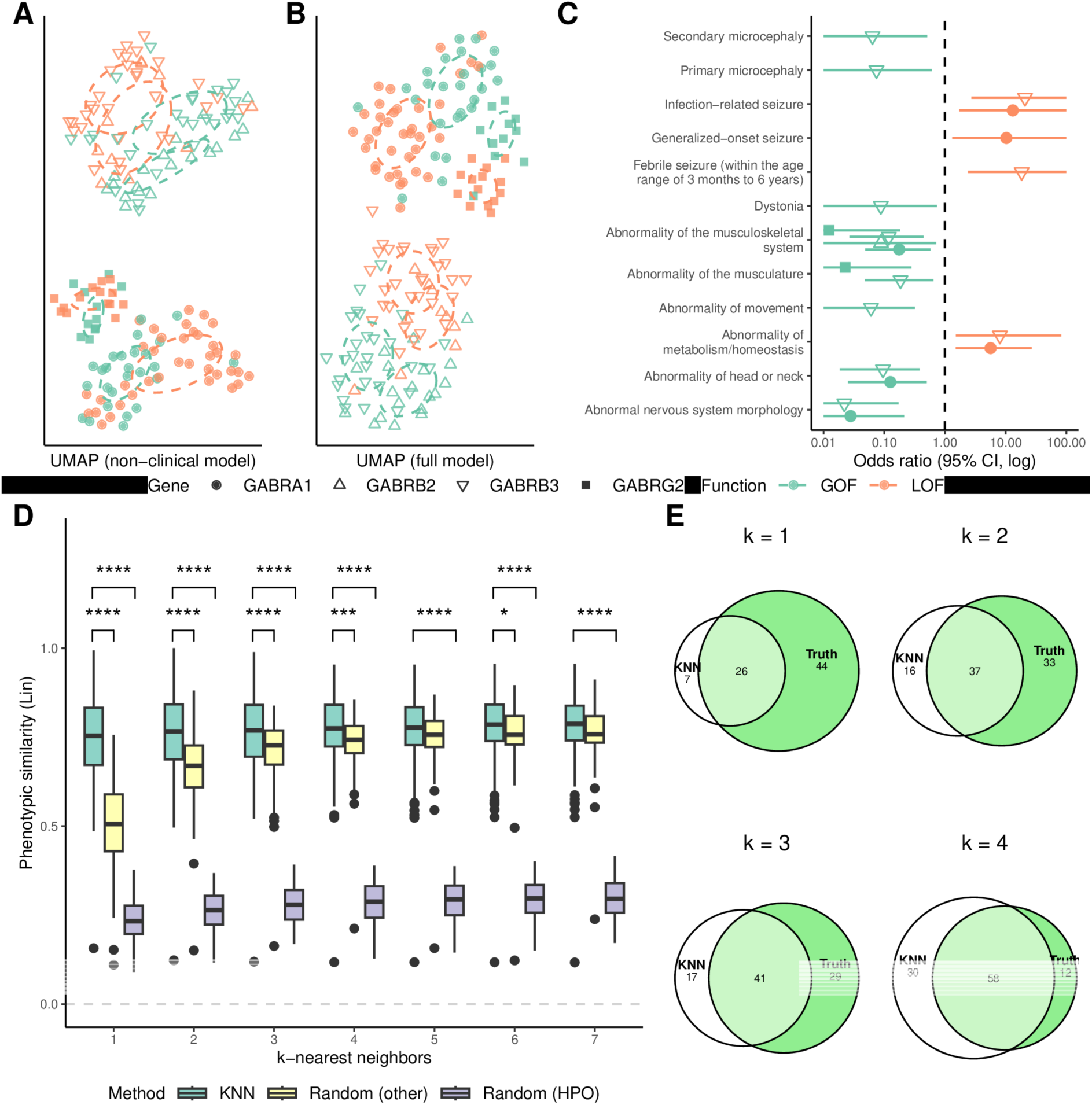
GENTLY may provide further clinical insight into GABA_A_-receptor related disorders. **(A)** UMAP (Uniform Manifold Approximation and Projection) plot of the feature space of the model with no clinical features. Note that genes and functional effects are visually separable. Normal distributions are shown as dashed circles. **(B)** UMAP plot of the full model, where the inclusion of clinical features improves separability and thus prediction of functional effects. **(C)** Clinical spectrum of GABA_A_ variants by functional effect. Forest plot of relative enrichment (Fisher’s exact test) of HPO terms across functional subgroups and genes. **(D)** The feature space of the model with no clinical features (A) can be used to estimate the likely phenotype with a k-nearest neighbour approach. Two-sided t-test. Random (other) corresponds to phenotypes based on any non-neighbour variants. Random (HPO) corresponds to phenotypes based on random human phenotype ontology (HPO) terms sampled from the ontology. Baseline estimates were repeated 10-fold. Reconstructed phenotypes are compared to the clinically observed phenotype using the Lin similarity measure (range 0-1, with higher values denoting higher phenotypic similarity). **(E)** Venn diagram demonstrating the overlap of reconstructed versus observed HPO terms for different values of k in k-nearest neighbours. The variant *GABRA1* p.V270I is shown as an example. Significance: *p < 0.05, **p < 0.01, ***p<0.001, ****p<0.0001.

To explore the feasibility of phenotype prediction from variant-level structural information, we employed a *k*-nearest neighbour approach: For each variant, we found the *k* nearest variants in the feature space of the model with no clinical features (Fig. 5A) and compared the set of their HPO terms to the variant of interest. Using this approach, meaningful variant phenotypes could be predicted (median Lin similarity 0.754 IQR 0.161 versus random baseline median 0.506 IQR 0.160; two-sided t-test: *p* < 0.0001; Lin similarity range 0-1, with higher values denoting higher phenotypic similarity; Fig. 5D). For example, 26/70 (37.1%) to 58/70 (82.9%) of clinical characteristics of the variant *GABRA1* p.V270I could be reconstructed using *k* = 1 to 4 nearest variants in feature space (Fig. 5E). Thus, the variant representation of our model could be used to infer likely clinical characteristics, pending further validation.

## Discussion

In this study, we used comprehensive clinical and electrophysiological data from 505 affected individuals with 272 GABA_A_ receptor variants to train a machine learning model to predict variant functional effects (GOF or LOF). This model was then externally validated on a further 197 affected individuals with 138 GABA_A_ receptors variants. We found that our model outperforms current state-of-the-art genome-wide functional effect predictions algorithms by a wide margin. This was expected: First, previous studies have demonstrated that gene-specific model design can make efficient use of expert knowledge by explicit representation of underlying mechanisms (e.g. gene family taxonomy, specific binding sites and motifs, pore and membrane geometry) which the genome-wide deep learning models may not represent well.^26–30^ Further, this biologically grounded design makes our model more plausible and robust as demonstrated by conformation to expert-based structure-function hypotheses. Second, genome-wide predictors work on more general ‘genomic’ definitions of GOF (e.g. hypermorphs, where protein activity is increased, and neomorphs, where novel functions are acquired) and LOF (e.g. destabilisation of protein folding).^66,67^ These assumptions do not seem to reflect the very specific GOF/LOF mechanisms observed in the electrophysiological characterization of ion channels and transporters. Thus, specialised prediction models are needed.

Previous models were limited by data availability as electrophysiological data is time-consuming and expensive to acquire, which has resulted in the use of data from non-human channels or inference of labels from observed phenotypes.^26,29^ Accordingly, validation datasets were small and actual model performance was largely uncertain. Here, we utilized the largest family-specific clinical and electrophysiological dataset to-date. We aimed to minimize noise or uncertainty in our dataset, improving predictive power and reliability: First, clinical data were harmonised by independent assessment from experienced clinicians and further standardized using ontological reasoning with HPO terms.^37^ Second, all electrophysiological data was recorded at the same laboratory using receptor assemblies that directly reflect heteropentamer composition in heterozygous carriers.^68,69^ Together, these steps ensured data quality and resulted in a highly calibrated model where prediction confidence directly correlated with quantitative measurements for the severity of the electrophysiological changes.

Functional prediction algorithms are designed to address a key translational gap in neurodevelopmental disorders: While the increasing utilization of next-generation sequencing has outpaced the availability of direct experimental evidence, ongoing natural history and precision medicine trials (including both specific agonists/antagonists, and antisense oligonucleotide or gene therapies) in rare disease require functional evidence for patient identification and stratification (e.g. NCT06872125, NCT05419492). Currently, *in-vitro* electrophysiology remains the gold standard and no functional prediction algorithms have been approved for clinical use. However, if we want to pave the way for their implementation, we must address two questions: First, how does the model respond when provided with a (likely) benign variant from the general population? This represents a common use case where the variant of interest has initially been misclassified. Here, we provide a score range common for population variants that should prompt variant reassessment (probability of GOF: 0.208<x<0.283; probability of LOF: 0.717<x<0.792), thus minimizing risk. Further, we provide top score percentiles for high-confidence predictions (probability of LOF: >0.928; probability of GOF >0.850), and ACMG-calibrated thresholds. Second, does our model offer additional insight into case variants which may prompt reclassification? We note that the ACMG criterion PP3 (computational and predictive data) currently only applies to a set of well-calibrated predictors (e.g. REVEL).^63,70^ Similarly, ACMG criterion PS3 (functional data) is reserved for well-established functional assays.^71^ Yet, our model may provide complementary evidence. Future use will require additional prospective validation and integration in multimodal evidence frameworks.

Regarding limitations, we note that the performance of our full model deteriorated on external validation in two specific genes not observed during training (e.g. *GABRA3, GABRD*). This was not unexpected and rather reflects that the model learnt valid information from the gene family: First, LOF in either gene has not yet been clearly linked to epilepsy and thus does not align with the established genotype-phenotype correlations.^17,72^ Second, the genotype of *GABRA3* is an outlier as it is the only disease-associated X-linked gene in the family.^11^ Third, *GABRD* has rather different criteria for functional changes.^17^ Our recommendation would be to first establish gene-disease validity (i.e. not assuming causality for genes not yet linked to human disease), then assessing variant pathogenicity using ACMG criteria, before using the model. We would suggest using the structural model with the score thresholds provided above, complemented by expert assessment based on known genotype-phenotype correlations, including the clinical decision tree we have provided (Supplementary Fig. 2).^13,14^

Some potential bias from circular learning applies: *(i)* clinical assessment of the individuals in our study may have been biased by previously observed genotype-phenotype correlations; *(ii)* continuous labels (e.g. ΔpEC50) were partially used to assign categorical labels (GOF/LOF). However, these were only part of the full electrophysiological characterization and were done independently while the added value lies in the correlation with prediction confidence beyond a simple cut-off value; *(iii)* previous experimental data used during training indirectly informed knowledge on structure-function relationships. We consider it to still be useful that the model conforms to these observations, making expert biophysicist knowledge available to clinical users. In summary, we consider these possible biases to be acceptable as they represent current expert knowledge on GABA_A_ receptor subunits. Importantly, the model still performed well on external validation on a set of variants that were recorded after the training data. Lastly, we consider our approach to phenotypic learning and prediction to be exploratory pending larger cohorts and methodological refinement similar to that of the *SCN1A*-Dravet model.^65^

## Conclusion

Functional prediction of GABA_A_ receptor subunit variants is feasible, highly accurate, biologically plausible, and clinically meaningful. Our approach brings together methodological advances in machine learning with recent studies on the ever-expanding spectrum of GABA_A_ receptor related disorders. Pending further validation, variant functional effect prediction algorithms may facilitate timely diagnosis, inform genetic counselling, and enable candidate precision therapies including patient identification and stratification for clinical studies.

## Acknowledgements

We sincerely thank the patients, caregivers, families and patient advocacy groups, including CURE GABA-A, for their participation and support.

## Supplementary material

Pending full publication.

## References

1. Rossi A, Lin SXN, Absalom NL, et al. Phenotypic Spectrum in Individuals With Pathogenic GABRG2 Loss- and Gain-of-Function Variants. Neurology. 2025;105(2):e213644. doi:10.1212/WNL.0000000000213644

2. Baulac S, Huberfeld G, Gourfinkel-An I, et al. First genetic evidence of GABA(A) receptor dysfunction in epilepsy: a mutation in the gamma2-subunit gene. Nat Genet. 2001;28(1):46–48. doi:10.1038/ng0501-46

3. Epi4K Consortium, Epilepsy Phenome/Genome Project, Allen AS, et al. De novo mutations in epileptic encephalopathies. Nature. 2013;501(7466):217–221. doi:10.1038/nature12439

4. Absalom NL, Lin SXN, Liao VWY, et al. GABAA receptors in epilepsy: Elucidating phenotypic divergence through functional analysis of genetic variants. J Neurochem. 2024;168(12):3831–3852. doi:10.1111/jnc.15932

5. May P, Girard S, Harrer M, et al. Rare coding variants in genes encoding GABAA receptors in genetic generalised epilepsies: an exome-based case-control study. Lancet Neurol. 2018;17(8):699–708. doi:10.1016/S1474-4422(18)30215-1

6. Møller RS, Wuttke TV, Helbig I, et al. Mutations in GABRB3: From febrile seizures to epileptic encephalopathies. Neurology. 2017;88(5):483–492. doi:10.1212/WNL.0000000000003565

7. Hernandez CC, XiangWei W, Hu N, et al. Altered inhibitory synapses in de novo GABRA5 and GABRA1 mutations associated with early onset epileptic encephalopathies. Brain. 2019;142(7):1938–1954. doi:10.1093/brain/awz123

8. Katzner S, Busse L, Carandini M. GABAA inhibition controls response gain in visual cortex. J Neurosci. 2011;31(16):5931–5941. doi:10.1523/JNEUROSCI.5753-10.2011

9. Cossette P, Liu L, Brisebois K, et al. Mutation of GABRA1 in an autosomal dominant form of juvenile myoclonic epilepsy. Nat Genet. 2002;31(2):184–189. doi:10.1038/ng885

10. Butler KM, Moody OA, Schuler E, et al. De novo variants in GABRA2 and GABRA5 alter receptor function and contribute to early-onset epilepsy. Brain. 2018;141(8):2392–2405. doi:10.1093/brain/awy171

11. Niturad CE, Lev D, Kalscheuer VM, et al. Rare GABRA3 variants are associated with epileptic seizures, encephalopathy and dysmorphic features. Brain. 2017;140(11):2879–2894. doi:10.1093/brain/awx236

12. Millevert C, Kan ASH, Hanke M, et al. The genetic and phenotypic spectrum of GABRB1-related disorders. Brain. Published online June 5, 2025:awaf213. doi:10.1093/brain/awaf213

13. Mohammadi NA, Ahring PK, Yu Liao VW, et al. Distinct neurodevelopmental and epileptic phenotypes associated with gain- and loss-of-function GABRB2 variants. eBioMedicine. 2024;106:105236. doi:10.1016/j.ebiom.2024.105236

14. Absalom NL, Liao VWY, Johannesen KMH, et al. Gain-of-function and loss-of-function GABRB3 variants lead to distinct clinical phenotypes in patients with developmental and epileptic encephalopathies. Nat Commun. 2022;13(1):1822. doi:10.1038/s41467-022-29280-x

15. Absalom NL, Liao VWY, Kothur K, et al. Gain-of-function GABRB3 variants identified in vigabatrin-hypersensitive epileptic encephalopathies. Brain Commun. 2020;2(2):fcaa162. doi:10.1093/braincomms/fcaa162

16. Kananura C, Haug K, Sander T, et al. A Splice-Site Mutation in GABRG2 Associated With Childhood Absence Epilepsy and Febrile Convulsions. Archives of Neurology. 2002;59(7):1137–1141. doi:10.1001/archneur.59.7.1137

17. Ahring PK, Liao VWY, Gardella E, et al. Gain-of-function variants in GABRD reveal a novel pathway for neurodevelopmental disorders and epilepsy. Brain. 2022;145(4):1299–1309. doi:10.1093/brain/awab391

18. Beltrán-Corbellini Á, Aledo-Serrano Á, Møller RS, et al. Epilepsy Genetics and Precision Medicine in Adults: A New Landscape for Developmental and Epileptic Encephalopathies. Front Neurol. 2022;13:777115. doi:10.3389/fneur.2022.777115

19. Knowles JK, Helbig I, Metcalf CS, et al. Precision medicine for genetic epilepsy on the horizon: Recent advances, present challenges, and suggestions for continued progress. Epilepsia. 2022;63(10):2461–2475. doi:10.1111/epi.17332

20. Striano P, Minassian BA. From Genetic Testing to Precision Medicine in Epilepsy. Neurotherapeutics. 2020;17(2):609–615. doi:10.1007/s13311-020-00835-4

21. Chen E, Facio FM, Aradhya KW, et al. Rates and Classification of Variants of Uncertain Significance in Hereditary Disease Genetic Testing. JAMA Netw Open. 2023;6(10):e2339571. doi:10.1001/jamanetworkopen.2023.39571

22. Fowler DM, Rehm HL. Will variants of uncertain significance still exist in 2030? The American Journal of Human Genetics. 2024;111(1):5–10. doi:10.1016/j.ajhg.2023.11.005

23. eClinicalMedicine. Whole-genome sequencing for every newborn in the UK: promise and practicalities. eClinicalMedicine. 2025;85:103387. doi:10.1016/j.eclinm.2025.103387

24. Bauskis A, Strange C, Molster C, Fisher C. The diagnostic odyssey: insights from parents of children living with an undiagnosed condition. Orphanet Journal of Rare Diseases. 2022;17(1):233. doi:10.1186/s13023-022-02358-x

25. Faye F, Crocione C, Anido de Peña R, et al. Time to diagnosis and determinants of diagnostic delays of people living with a rare disease: results of a Rare Barometer retrospective patient survey. Eur J Hum Genet. 2024;32(9):1116–1126. doi:10.1038/s41431-024-01604-z

26. Heyne HO, Baez-Nieto D, Iqbal S, et al. Predicting functional effects of missense variants in voltage-gated sodium and calcium channels. Sci Transl Med. 2020;12(556):eaay6848. doi:10.1126/scitranslmed.aay6848

27. Brunklaus A, Feng T, Brünger T, et al. Gene variant effects across sodium channelopathies predict function and guide precision therapy. Brain. 2022;145(12):4275–4286. doi:10.1093/brain/awac006

28. Montanucci L, Brünger T, Bhattarai N, et al. Ligand distances as key predictors of pathogenicity and function in NMDA receptors. Hum Mol Genet. Published online November 13, 2024:ddae156. doi:10.1093/hmg/ddae156

29. Boßelmann CM, Hedrich UBS, Müller P, et al. Predicting the functional effects of voltage-gated potassium channel missense variants with multi-task learning. EBioMedicine. 2022;81:104115. doi:10.1016/j.ebiom.2022.104115

30. Boßelmann CM, Hedrich UBS, Lerche H, Pfeifer N. Predicting functional effects of ion channel variants using new phenotypic machine learning methods. PLOS Computational Biology. 2023;19(3):e1010959. doi:10.1371/journal.pcbi.1010959

31. Kan ASH, Kusay AS, Mohammadi NA, et al. Understanding paralogous epilepsy–associated GABAA receptor variants: Clinical implications, mechanisms, and potential pitfalls. Proceedings of the National Academy of Sciences. 2024;121(50):e2413011121. doi:10.1073/pnas.2413011121

32. Brünger T, Pérez-Palma E, Montanucci L, et al. Conserved patterns across ion channels correlate with variant pathogenicity and clinical phenotypes. Brain. 2023;146(3):923–934. doi:10.1093/brain/awac305

33. Brünger T, Ivaniuk A, Pérez-Palma E, et al. Conserved missense variant pathogenicity and correlated phenotypes across paralogous genes. Genome Biol. 2025;26(1):197. doi:10.1186/s13059-025-03663-x

34. Absalom NL, El-Kamand S, Chua HC, Ahring PK. Follow the allosteric transitions to predict variant pathogenicity: a channel-specific approach. Brain. 2024;147(5):e37–e40. doi:10.1093/brain/awae008

35. Richards S, Aziz N, Bale S, et al. Standards and guidelines for the interpretation of sequence variants: a joint consensus recommendation of the American College of Medical Genetics and Genomics and the Association for Molecular Pathology. Genet Med. 2015;17(5):405–424. doi:10.1038/gim.2015.30

36. Harris PA, Taylor R, Minor BL, et al. The REDCap consortium: Building an international community of software platform partners. J Biomed Inform. 2019;95:103208. doi:10.1016/j.jbi.2019.103208

37. Gargano MA, Matentzoglu N, Coleman B, et al. The Human Phenotype Ontology in 2024: phenotypes around the world. Nucleic Acids Res. 2024;52(D1):D1333–D1346. doi:10.1093/nar/gkad1005

38. Widmer C, Leiva J, Altun Y, Rätsch G. Leveraging Sequence Classification by Taxonomy-Based Multitask Learning. In: Berger B, ed. Research in Computational Molecular Biology. Springer; 2010:522–534. doi:10.1007/978-3-642-12683-3_34

39. Edgar RC. MUSCLE: a multiple sequence alignment method with reduced time and space complexity. BMC Bioinformatics. 2004;5(1):113. doi:10.1186/1471-2105-5-113

40. Schölkopf B, Smola AJ. Learning with Kernels: Support Vector Machines, Regularization, Optimization, and Beyond. MIT Press; 2002.

41. Jumper J, Evans R, Pritzel A, et al. Highly accurate protein structure prediction with AlphaFold. Nature. 2021;596(7873):583–589. doi:10.1038/s41586-021-03819-2

42. Wilson CM, Li K, Yu X, Kuan PF, Wang X. Multiple-kernel learning for genomic data mining and prediction. BMC Bioinformatics. 2019;20(1):426. doi:10.1186/s12859-019-2992-1

43. Xu Z, Jin R, Yang H, King I, Lyu MR. Simple and efficient multiple kernel learning by group lasso. In: Proceedings of the 27th International Conference on International Conference on Machine Learning. ICML’10. Omnipress; 2010:1175–1182.

44. Rakotomamonjy A, Bach FR, Canu S, Grandvalet Y. SimpleMKL. Journal of Machine Learning Research. 2008;9(83):2491–2521.

45. Kloft M, Brefeld U, Laskov P, Müller KR, Zien A, Sonnenburg S. Efficient and Accurate Lp-Norm Multiple Kernel Learning. In: Advances in Neural Information Processing Systems. Vol 22. Curran Associates, Inc.; 2009. Accessed October 13, 2025. https://proceedings.neurips.cc/paper_files/paper/2009/hash/d516b13671a4179d9b7b458a6ebdeb92-Abstract.html

46. Wainer J, Cawley G. Nested cross-validation when selecting classifiers is overzealous for most practical applications. Expert Systems with Applications. 2021;182:115222. doi:10.1016/j.eswa.2021.115222

47. Platt J. Probabilistic Outputs for Support Vector Machines and Comparisons to Regularized Likelihood Methods. Adv Large Margin Classif. 2000;10.

48. Stein D, Kars ME, Wu Y, et al. Genome-wide prediction of pathogenic gain- and loss-of-function variants from ensemble learning of a diverse feature set. Genome Med. 2023;15(1):103. doi:10.1186/s13073-023-01261-9

49. Brixi G, Durrant MG, Ku J, et al. Genome modeling and design across all domains of life with Evo 2. bioRxiv. Preprint posted online February 21, 2025:2025.02.18.638918. doi:10.1101/2025.02.18.638918

50. Sun KY, Bai X, Chen S, et al. A deep catalog of protein-coding variation in 985,830 individuals. bioRxiv. Published online November 2, 2023:2023.05.09.539329. doi:10.1101/2023.05.09.539329

51. Karczewski KJ, Francioli LC, Tiao G, et al. The mutational constraint spectrum quantified from variation in 141,456 humans. Nature. 2020;581(7809):434–443. doi:10.1038/s41586-020-2308-7

52. Landrum MJ, Lee JM, Benson M, et al. ClinVar: improving access to variant interpretations and supporting evidence. Nucleic Acids Res. 2018;46(D1):D1062–D1067. doi:10.1093/nar/gkx1153

53. Friendly M, Monette G, Fox J. Elliptical Insights: Understanding Statistical Methods through Elliptical Geometry. Statistical Science. 2013;28(1):1–39. doi:10.1214/12-STS402

54. Cherkassky V, Dhar S. Simple Method for Interpretation of High-Dimensional Nonlinear SVM Classification Models. In: 2010:267–272.

55. Greene D, Richardson S, Turro E. ontologyX: a suite of R packages for working with ontological data. Bioinformatics. 2017;33(7):1104–1106. doi:10.1093/bioinformatics/btw763

56. Karatzoglou A, Smola A, Hornik K, Zeileis A. kernlab - An S4 Package for Kernel Methods in R. Journal of Statistical Software. 2004;11:1–20. doi:10.18637/jss.v011.i09

57. Wickham H, Averick M, Bryan J, et al. Welcome to the Tidyverse. Journal of Open Source Software. 2019;4(43):1686. doi:10.21105/joss.01686

58. Frazer J, Notin P, Dias M, et al. Disease variant prediction with deep generative models of evolutionary data. Nature. 2021;599(7883):91–95. doi:10.1038/s41586-021-04043-8

59. Montanucci L, Brünger T, Boßelmann CM, et al. Evaluating novel in silico tools for accurate pathogenicity classification in epilepsy-associated genetic missense variants. Epilepsia. 2024;65(12):3655–3663. doi:10.1111/epi.18155

60. SoRelle JA, Pascual JM, Gotway G, Park JY. Assessment of Interlaboratory Variation in the Interpretation of Genomic Test Results in Patients With Epilepsy. JAMA Netw Open. 2020;3(4):e203812. doi:10.1001/jamanetworkopen.2020.3812

61. Gunning AC, Fryer V, Fasham J, et al. Assessing performance of pathogenicity predictors using clinically relevant variant datasets. J Med Genet. 2021;58(8):547–555. doi:10.1136/jmedgenet-2020-107003

62. Whiffin N, Minikel E, Walsh R, et al. Using high-resolution variant frequencies to empower clinical genome interpretation. Genet Med. 2017;19(10):1151–1158. doi:10.1038/gim.2017.26

63. Pejaver V, Byrne AB, Feng BJ, et al. Calibration of computational tools for missense variant pathogenicity classification and ClinGen recommendations for PP3/BP4 criteria. Am J Hum Genet. 2022;109(12):2163–2177. doi:10.1016/j.ajhg.2022.10.013

64. Bergquist T, Stenton SL, Nadeau EAW, et al. Calibration of additional computational tools expands ClinGen recommendation options for variant classification with PP3/BP4 criteria. Genet Med. 2025;27(6):101402. doi:10.1016/j.gim.2025.101402

65. Brunklaus A, Pérez-Palma E, Ghanty I, et al. Development and Validation of a Prediction Model for Early Diagnosis of SCN1A-Related Epilepsies. Neurology. 2022;98(11):e1163–e1174. doi:10.1212/WNL.0000000000200028

66. Gerasimavicius L, Livesey BJ, Marsh JA. Loss-of-function, gain-of-function and dominant-negative mutations have profoundly different effects on protein structure. Nat Commun. 2022;13(1):3895. doi:10.1038/s41467-022-31686-6

67. Badonyi M, Marsh JA. Prevalence of loss-of-function, gain-of-function and dominant-negative mechanisms across genetic disease phenotypes. Nat Commun. 2025;16(1):8392. doi:10.1038/s41467-025-63234-3

68. Liao VWY, Chua HC, Kowal NM, Chebib M, Balle T, Ahring PK. Concatenated γ-aminobutyric acid type A receptors revisited: Finding order in chaos. J Gen Physiol. 2019;151(6):798–819. doi:10.1085/jgp.201812133

69. Liao VWY, Chebib M, Ahring PK. Efficient expression of concatenated α1β2δ and α1β3δ GABAA receptors, their pharmacology and stoichiometry. Br J Pharmacol. 2021;178(7):1556–1573. doi:10.1111/bph.15380

70. Bergquist T, Stenton SL, Nadeau EAW, et al. Calibration of additional computational tools expands ClinGen recommendation options for variant classification with PP3/BP4 criteria. bioRxiv. Published online September 21, 2024:2024.09.17.611902. doi:10.1101/2024.09.17.611902

71. Brnich SE, Abou Tayoun AN, Couch FJ, et al. Recommendations for application of the functional evidence PS3/BS3 criterion using the ACMG/AMP sequence variant interpretation framework. Genome Medicine. 2019;12(1):3. doi:10.1186/s13073-019-0690-2

72. Johannesen KM, Aung KP, Liao VW, et al. Functional consequence of pathogenic GABRA3 variants determines whether X-linked inheritance is dominant or recessive. J Clin Invest. Published online November 25, 2025:e189830. doi:10.1172/JCI189830

